# On the predictability of postoperative complications for cancer patients: a Portuguese cohort study

**DOI:** 10.1101/2021.03.27.21254473

**Authors:** Daniel Gonçalves, Rui Henriques, Lúcio Lara Santos, Rafael S Costa

## Abstract

Postoperative complications following cancer surgeries are still hard to predict despite the historical efforts towards the creation of standard clinical risk scores. The differences among score calculators, contribute for the creation of highly specialized tools, with poor reusability in foreign contexts, resulting in larger prediction errors in clinical practice.

This work aims to predict postoperative complications risk for cancer patients, offering two major contributions. First, to develop and evaluate a machine learning-based risk score, specific for the Portuguese population using a retrospective cohort of 847 cancer patients undergoing surgery between 2016 and 2018, predicting 4 outcomes of interest: i) existence of postoperative complications, ii) severity level of complications, iii) number of days in the Intermediate Care Unit (ICU), and iv) postoperative mortality within 1 year. An additional cohort of 137 cancer patients was used to validate the models. Second, to support the study with relevant findings and improve the interpretability of predictive models.

In order to achieve these objectives, a robust methodology for the learning of risk predictors is proposed, offering new perspectives and insights into the clinical decision process. For postoperative complications the mean Receiver Operating Characteristic Curve (AUC) was 0.69, for complications’ severity mean AUC was 0.65, for the days in the ICU the Mean Absolute Error (MAE) was 1.07 days, and for one-year postoperative mortality the mean AUC was 0.74, calculated on the development cohort.

In this study, risk predictive models which may help guide physicians at estimating cancer patient’s risk of developing surgical complications were developed. Additionally, a web-based decision support system is further provided to this end.

## Introduction

Cancer is a major health problem worldwide and it is among the leading death causes of the 21^st^ century. There are at least two battlefronts in reducing deaths associated to cancer, those resulting from direct consequences of the disease, and those occurring due to complications from surgery treatment [1]. Surgical complications contribute to lower survival probability and, in certain types of cancer, to aggravate the recurrence rate [1, 2, 3, 4]. The outcome of such surgeries is still widely unpredictable due to the large number of factors involved. In an attempt to facilitate perioperative risk assessment for the selection of patients benefiting from surgery, a variety of traditional scoring systems incorporating several risk factors have been developed [5].

From a clinical perspective, the traditional risk scores (e.g., P-POSSUM [6], ARISCAT [7] and ACS score [8]) are determinant in choosing the course of actions, such as prehabilitation or supportive measures, to be taken during the preoperative, intraoperative and postoperative periods [5]. Nevertheless, it is becoming clear that they are not adequate to support clinical decisions, particularly in the geriatric population, as predicted and observed rates often significantly differ [9].

Recently, machine learning (ML) models for surgical outcomes prediction have been proposed. ML comprises algorithms that can learn from a set of data and improve on their own, allowing for more accurate predictions [10]. For instance, Wang et al. [11] proposed several ML models to predict 5-year mortality in a bladder cancer cohort. The study used clinical and histopathological data from 117 patients, and achieved 80% accuracy using a Regularized Extreme Learning Machine, alongside Neural Networks, Naive Bayes, k-Nearest Neighbors and Support Vector Machines (SVM). More recently, Corey et al. [12] explored ML methods to identify high-risk surgical patients from a local institution using electronic health record data. Focusing several specific complication outcomes, this study establishes a threshold of 15% on any given outcome to define a patient as high-risk. The sensitivity and specificity were 76%, resulting from a set of ML models. Another example is the study conducted by Lee [13] where deep neural network models were successfully used to classify the risks of postoperative mortality, acute kidney injury, and reintubation, outperforming Logistic Regression, ASA [14] and the Surgical Apgar [15] scores. To this end, readily available intraoperative variables were utilized such as physiological waveforms as well as intermittent vital sign values, lab values, and ventilator settings. Furthermore, this study also employs Generalized Additive Neural Networks as a means to extract feature importance information and enhance the interpretability of otherwise black-box Neural Networks.

Despite the inherent potentialities of ongoing efforts, the exiting postoperative risk prediction studies in the cancer field are limited by the size of available hospital records, the lack of systematic evaluation of different models and no one comprehensively targets the Portuguese population. Identification of reliable prognostic factors, representative of our own patient population, may help clinicians not only to accurately select patients eligible for surgery, but also to identify high-risk patients that may benefit from individualized optimization with multimodal prehabilitation interventions. There is thus an urgent need to improve perioperative risk assessment to reduce the growing postoperative burden among patients who undergo surgery for cancer.

This work assesses the predictability of four major postoperative outcomes: i) existence of postoperative complications, ii) the severity of said complications, iii) the number of days in the ICU, and the iv) death within 1 year after surgery, in cancer patients. In this context, it offers two major contributions. First, a robust methodology for the prognostication of oncological surgeries. Secondly, this work further establishes principles to support the study of this disease and surgical prognostication, either by finding relevant variables, or improving the interpretability of these models.

This work is structured as follows. Methods section introduces the methodology used to conduct this study, covering the dataset, preprocessing, predictive models, outcomes, result optimization, assessment and validation. Results and Discussion sections present the major findings of this work. Finally, some concluding remarks and limitations are identified.

## Methods

### Dataset

The data derives from a single-center retrospective cohort study of cancer patients that have undertaken surgery at the Portuguese Institute of Oncology, Porto, Portugal (IPO-Porto), and were monitored from 2016 to 2018. The IPO-Porto Ethics Committee approved (CES IPO:91/019) the analysis of the anonymized data. The collected cohort of 847 patients contains information pertaining to the demographic and physiological data, cancer location, histopathological determinants, traditional risk score variables (from P-Possum [6], ACS NSQIP [8], ARISCAT [7]), surgical procedures and outcomes of interest. From a total of 136 routinely-collected variables, only 62 are available before surgery and therefore usable as input for a risk score. There are 20 binary variables, 20 ordinal, 10 categorical, 5 numeric, 2 in date format and 5 pure text variables (see Table 5 in Appendix C). The surgeries type were mainly elective, with only 11% of the procedures corresponding to emergencies. Figure 1 displays the summary of the cohort data. There are 11 types of cancer present and this disease incidence is mainly concentrated on older people, closer to the age of 65 (Fig.1-(a)). There are more men undergoing surgery and they are also more likely to develop postoperative complications than women (as shown in Fig.1-(b)). There are types of cancer more likely to complicate and more lethal than others (Fig.1-(e)), where Neurologic and Musculoskeletal cancers are portrayed as the most lethal types. The degree of Clavien-Dindo severity associated to the postoperative complications is similar across the different types (shown in Fig.1-(c)), where Neurologic cancers are portrayed as the type with more severe complications. The days in the ICU rarely exceed 2 to 3 days but can stretch as far as 2 weeks or more (Fig.1-(f)).

**Figure 1.**
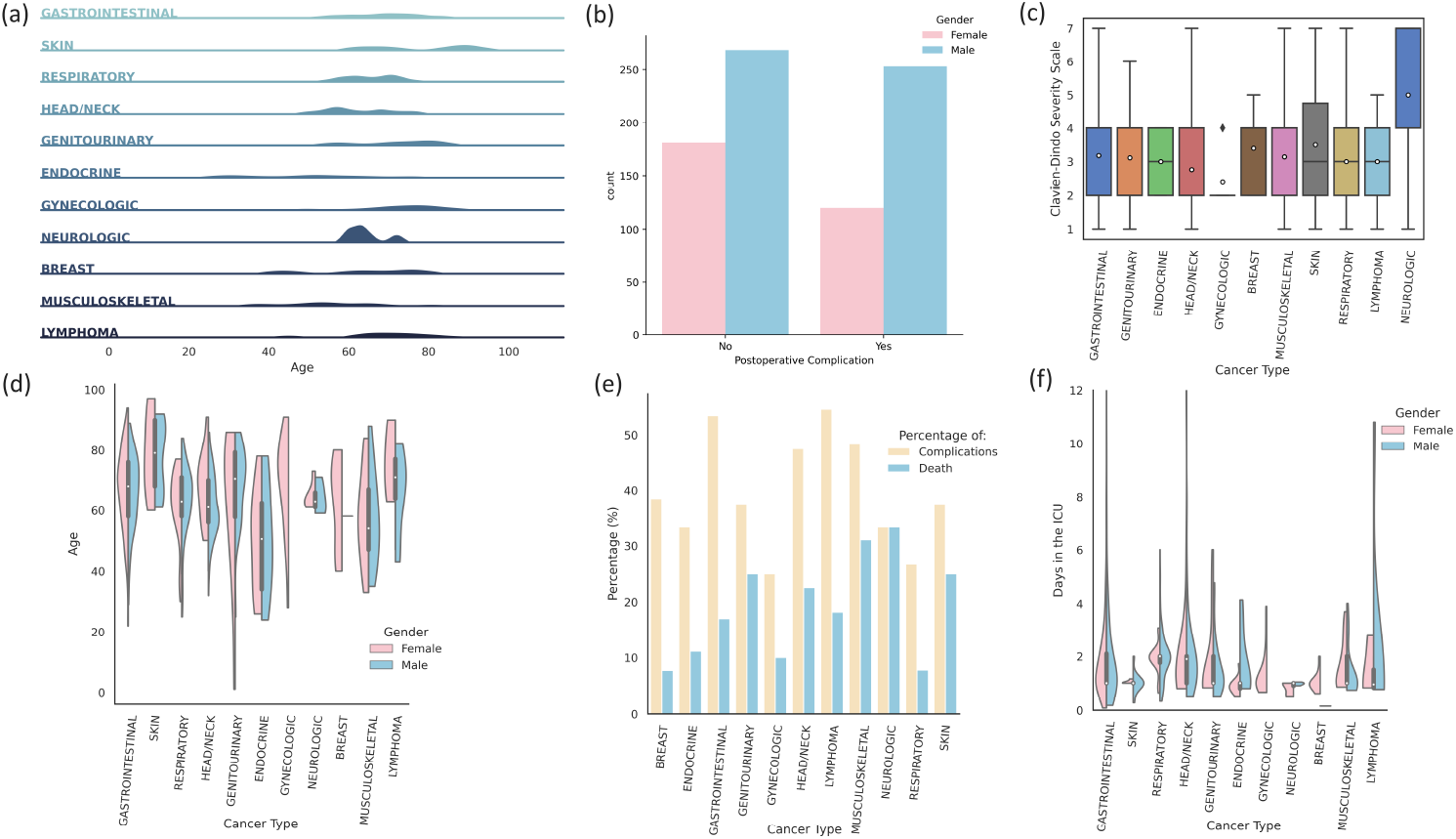
Cohort data overview: **(a)** Cancer type density plot according to patient age **(b)** Postoperative complications by gender **(c)** Complications’ severity by cancer type **(d)** Age distribution by gender and cancer type **(e)** Percentage of complications/deaths by cancer type **(f)** Distribution of days in the ICU by cancer type and gender.

### Data Preprocessing

The preprocessing is challenged by three major issues: missing values, mixed variables with non-identical distributions, imbalanced and sparse data (considering the variety of cancers and surgery types).

#### Missing Values

To minimize biases and predictive uncertainty, variables with high missing rate (*>*40%) were removed. In less extreme cases, and whenever classifiers are unable to handle missing data, missing values were imputed using an informed method based on the k-Nearest Neighbors algorithm [16], to help reduce the error introduced when dealing with missing values.

#### Categorical Variable Encoding

Categorical variables are commonly represented through a numeric encoding, which may not necessarily contain an implicit ordinal relationship. This quantitative or ordinal relationship might undesirably slip into the analysis. The simplest solution is to use a One-Hot encoder, consisting on splitting the categorical variable into a series of binary ones.

#### Resampling

To handle the observed imbalances on some of the outcomes for the target cohort study and avoid the bias of the classifiers towards the majority class, we apply a mixed strategy, combining synthetic oversampling with Tomek Links informed undersampling, as proposed by He and Garcia [17].

#### Feature Scaling

Numeric variables are normalized to promote the learning of the algorithms that are affected by the magnitude of the different input variables, commonly resulting in wrongfully attributed relevance.

#### Feature Selection

To account for differences on the relevance of input variables for a given outcome, a restricted number of variables were selected, as depicted in Figure 2. Filter methods offer a *p*-value representing the probability that a variable is not correlated to an outcome. The *χ*^2^ test is used to measure correlation for categorical variables, when the output is also categorical. The ANOVA correlation coefficient is used to measure the correlation between categorical and numeric variables (it is not relevant which one is the dependent variable). Pearson’s correlation coefficient is used when both the independent and the dependent variables are numeric.

**Figure 2.**
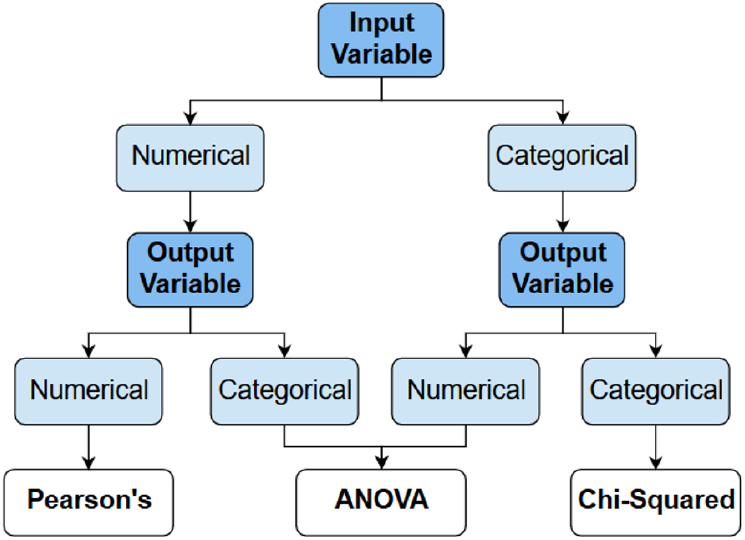
Scheme of the feature selection techniques used depending on the type of variables.

### Outcomes

In this work we attempt to address two main questions/outcomes: first, is a patient going to have postoperative complications? The postoperative complications were defined by any intervention (i.e. using diagnosis clinical codes) occurring within 90 days following the surgical procedure. Since the outcome is binary, a classification approach is used, with a discrete and well defined set of labels to attribute to a certain patient.

Secondly, how severe is the complication? The Clavien-Dindo classification system [18] in 4 major grades (excluding death) was followed for the classification of surgical complications. For this outcome, a multi-class classification approach is followed.

The probability of death is a relevant indicator to estimate the existence of future complications, and also the viability of surgery for a certain patient. In this case, death might not be the result of postoperative complications exclusively, but rather a combination of factors. We conducted this outcomes as a classification problem with the objective of predicting one-year mortality.

The **number of days spent in the ICU** represents important information for medical and also hospital management reasons. Due to the continuous and purely numeric nature, regression models are used to this end.

### ML Models

We used a set of state-of-the-art supervised ML models, and assessed the predictive performance of all.

- The classifier-based prediction algorithms were: Naive Bayes (NB), k-Nearest Neighbours (kNN), Decision Trees (DT), Random Forests (RF), Support Vector Machines (SVM), Logistic Regression (LR), Multilayer Perceptron (MLP), XGBoost Classifier (XGB) and CatBoost Classifier (CBC);
- The regression-based prediction algorithms were: Linear Regression, Ridge Regression, Lasso Regression, SVM Regressor, Elastic Regression, k-Nearest Neighbours Regressor, Decision Tree Regressor, Random Forest Regressor, XGBoost Regressor, Partial Least Squares Regression (PLS), Multilayer Perceptron Regressor and CatBoost Regressor (CBR).

All the models’ implementations were performed using the Scikit-learn[19] Python package. For the XGBoost[20] and CatBoost[21] algorithms two independent packages were used.

### Hyperparameter Optimization

The hyperparameters of the models were selected, using informed search methods. Bayesian optimization [22] associates a probability distribution to the hyperparameters tested, making the search faster than exhaustive approaches. Two objective functions were used:

- Regression models are optimized with respect to the Root Mean Squared Error (RMSE);
- Classification models are optimized to maximize their F1-Score (the harmonic mean of precision and recall).

### Development Process

The development process was performed in two phases: training and testing using cross-validation (within the primary dataset); independent validation (training with the primary dataset and testing on the secondary external one). Both begin by preprocessing the input data before feeding it to the models, either to learn or directly predict the outputs. The difference is that in the first phase there is an intermediate step for hyperparameter optimization and in the second phase such parameters are already available. All this was implemented in Python and the models used are presented in the previous sub-section. All the code written to process and analyze the data can be found in GitHub (https://github.com/danielmg97/cancer-prognostication-iposcore).

After models selection and optimization, a web-based application for use in the clinical context was built using the Dash library in Python (https://iposcore.herokuapp.com/).

### Performance Metrics and Validation

#### Classification Evaluation Metrics

The discrete nature of classifiers allows for simple evaluation. Given the imbalanced nature of data, accuracy is complemented with other metrics, like recall/sensitivity. The Receiver Operating Characteristic (ROC) curve can also be used to assess the model performance specifically as a measure of class separability. It is most commonly used in binary outcome settings but can be used for multi-class outcomes. In the latter, the AUC (Area Under the Curve) is more suitable and is employed in our assessment. The F1-score [23] combines precision and recall in a weighted average. This metric is the focus of our optimization efforts in order to guarantee the optimal sensitivity to every output class, even in multi-class settings where this measure is macro averaged. Cohen’s Kappa [24] is also used as a chance corrected standardized measure of agreement.

#### Regression Evaluation Metrics

In contrast with previous confusion-based metrics, residue-based scores are proposed to assess the predictability of numeric outcomes. RMSE is a quadratic scoring rule that also measures the average magnitude of the error. Since the errors are squared before they are averaged RMSE gives a larger weight to larger errors. The mean absolute error (MAE) measures the average magnitude of the errors on a set of predictions, complementing RMSE. Apart from checking the absolute fitment of the model, the Coefficient of Determination, or *R*^2^, is also used to assesses the fitness of the model to the available (training) data.

#### Model Validation

We used 10-fold cross-validation to assess the models’ ability to generalize into unseen data and also its performance variability, by testing in various sets of instances, allowing for more robust comparisons to be established.

#### External Validation

The models were externally validated on an independent cohort with 137 patient registries from the same hospital and had surgeries between January and October of 2019.

## Results

The importance of surgical risk stratification to drive interventions is well known. In this study, we analyzed the predictive performance of ML models for four major outcomes derived from our patient population, contributing to the improvement of cancer patients health, enhanced expectations management, and to the efficient management of hospital resources.

One of the constraints for the feasibility of this score’s application was the number of input variables. The number of overall inputs should not be greater than 20, as established by the clinicians in this study. To that end, the p-value threshold used in the feature selection process, as described previously, was fixed at 0.0002, resulting in an aggregation of 20 total variables used to calculate the different scores. The model selection process was performed according to specific target metrics: F1-Score for classification and RMSE for regression. Detailed information for all variables is presented in Table 5 (Appendix C). Results are gathered and discussed below.

### Postoperative Complications

The performance of the top 5 models for the postoperative complication outcomes are shown in Table 1. The results shown that it is possible to predict the presence of postoperative complications with 65% accuracy and 0.69 AUC by a Random Forest using only 8 input variables (see Table 5 - Appendix C), after the feature selection process (Figure 2). Other algorithms are able to achieve similar predictive performance, but are outperformed by the RF that can be a more easily interpretable solution upon individual tree analysis, when compared with alternatives such as the MLP.

**Table 1.** Top 5 models for the postoperative complications outcome, obtained through cross-validation inside the primary 847 patients dataset. (Mean *±* SD)

| Model | Kappa | Recall | AUC | F1-Score | Accuracy |
| --- | --- | --- | --- | --- | --- |
| RF | 0.293 ± 0.095 | 0.645 ± 0.081 | 0.691 ± 0.057 | 0.645 ± 0.046 | 0.652 ± 0.048 |
| MLP | 0.285 ± 0.096 | 0.642 ± 0.101 | 0.663 ± 0.053 | 0.641 ± 0.050 | 0.648 ± 0.048 |
| SVM | 0.282 ± 0.121 | 0.640 ± 0.098 | 0.676 ± 0.058 | 0.640 ± 0.060 | 0.646 ± 0.061 |
| CBC | 0.276 ± 0.109 | 0.636 ± 0.131 | 0.681 ± 0.064 | 0.635 ± 0.055 | 0.646 ± 0.053 |
| LR | 0.272 ± 0.086 | 0.634 ± 0.140 | 0.685 ± 0.056 | 0.632 ± 0.041 | 0.645 ± 0.044 |

Additionally, as proposed in our methodology, the models were validated in an independent set of 137 patients. The RF achieved an accuracy of 67% and an AUC of 0.71 (Appendix A), and the overall metrics achieve higher results, further supporting the generalization ability of our solution.

There are some studies aiming to predict the risk of any postoperative complication. Recently, Bihorac et al.[25] studied this outcome while addressing several specific complications, with AUC values ranging from 0.82 to 0.94, in a cohort of 51,457 patients. Corey et al. [12] also employed ML methods to predict a similar composite outcome, using a cohort of 66,370 patients, obtaining AUC values ranging from 0.75 and 0.92, a sensitivity of 0.78 and a specificity of 0.75. This is similar to our best risk models, and will potentially be helpful to complement medical prognosis for cancer patients undergoing surgery in the Portuguese hospitals.

Furthermore, IPO-Porto previously developed a simple Logistic Regression model, *MyIPOrisk-score* [26], based on the Age, Gender, P-Possum (Physiological) score and ACS NSQIP (serious complications) score to predict the probability of developing postoperative complications. This study was developed using 341 digestive cancer patients and obtained an AUC of 0.808 for the same set of patients. While we could not calculate the AUC (due to only having the binary output available), this traditional score performed inferiorly to the RF for the 137 independent validation patients, with accuracy = 0.613, F1-score = 0.101 and Cohen’s Kappa = 0.044.

### Severity of Complications

The complications’ severity was the second outcome of interest. Table 2 compares the predictive performance of the top 5 models. Overall, the predictability is in line with expectations for a 4 degree scale in a very imbalanced setting, with underrepresented grades. Being a harder prediction task, the feature selection process considered a higher amount of variables when compared with other outcomes, using 15 of the total 20 inputs (Table 5 - Appendix C). The best model in study is a RF, which is able to predict the output with an accuracy of 51% and 0.65 AUC.

**Table 2.** Top 5 models for the complication’s severity outcome, obtained through cross-validation inside the primary 847 patients dataset. (Mean *±* SD)

| Model | Kappa | Recall | AUC | F1-Score | Accuracy |
| --- | --- | --- | --- | --- | --- |
| RF | 0.225 ± 0.127 | 0.431 ± 0.164 | 0.651 ± 0.083 | 0.410 ± 0.093 | 0.506 ± 0.081 |
| CBC | 0.197 ± 0.098 | 0.430 ± 0.239 | 0.634 ± 0.089 | 0.377 ± 0.082 | 0.434 ± 0.071 |
| DT | 0.185 ± 0.118 | 0.388 ± 0.254 | 0.620 ± 0.094 | 0.368 ± 0.095 | 0.465 ± 0.083 |
| SVM | 0.157 ± 0.096 | 0.431 ± 0.243 | 0.642 ± 0.069 | 0.357 ± 0.062 | 0.393 ± 0.055 |
| XGB | 0.158 ± 0.128 | 0.424 ± 0.221 | 0.629 ± 0.062 | 0.354 ± 0.103 | 0.379 ± 0.091 |

In the independent validation set, the RF was able to predict the outcome with similar results, an accuracy of 61% and an AUC of 0.84 (Appendix A).

Burke et al. [27] targeted only grades IV and V (life-threatening and requiring intensive care unit management or death) of complications’ severity for 30 days after non-elective cholecystectomy. This study uses Logistic Regression to predict the risk level (low, medium or high) of surgical complications resulting in Clavien-Dindo IV and V grades. The results point to an AUC of 0.87 in the validation set. Being essentially a high severity complication risk predictor, the results can not be directly compared, but can be considered to be in line with the ones achieved in our study.

### Days in ICU

The prediction of days spent in the ICU is a difficult task given the typical short stays of 1 or 2 days, contrasting with a small percentage of patients with considerably longer stays. Although various transformations were used to attempt to minimize the effects of the skew in the data, the regressors are still inclined to predict lower values. Within the small improvements made along the development process, the models were able to achieve a MAE of approximately 1 day, by Ridge Regression (Table 3).

**Table 3.** Top 5 models for the days in the ICU outcome, obtained through cross-validation inside the primary 847 patients dataset. (Mean *±* SD)

| Model | MAE | RMSE | R<sup>2</sup> |
| --- | --- | --- | --- |
| Ridge | 1.071 ± 0.161 | 1.724 ± 0.436 | 0.042 ± 0.105 |
| Linear | 1.080 ± 0.157 | 1.729 ± 0.424 | 0.030 ± 0.122 |
| PLS | 1.079 ± 0.153 | 1.730 ± 0.420 | 0.029 ± 0.116 |
| MLPR | 1.075 ± 0.157 | 1.732 ± 0.426 | 0.029 ± 0.104 |
| RF | 1.077 ± 0.151 | 1.735 ± 0.428 | 0.027 ± 0.099 |

After the independent validation, the results of the best model remained identical, MAE of 1.07, RMSE of 1.77 and R^2^ of 0.07 (Appendix A).

The feature selection process indicated 7 relevant input variables (Table 5 - Appendix C), contributing for a lighter solution, which might mean a reduced data extraction effort for the hospital in the future. The 5 best models are presented in the Table 3.

Predicting the days in the intermediate care unit, can be an important part of predicting the length of hospital stays, allowing for better resource allocation. The studies found are generally aimed at predicting the total hospital stay length (including the various units where a patient might be) or at predicting the stays in Intensive Care Units. The number of days in ICU is typically short, as illustrated previously on Fig.1-(f), but these stays can stretch as far as 2 weeks. Our best models are able to predict this duration with an error close to 24 hours which could constitute critical information when longer stays are predicted, allowing for better resource allocation and to manage patient’s and doctor’s expectations.

### One-Year Mortality

This outcome shows good results when predicted by tree-based models, as shown in Table 4, achieving an accuracy of 85% and 0.74 AUC, by a Random Forest.

**Table 4.**
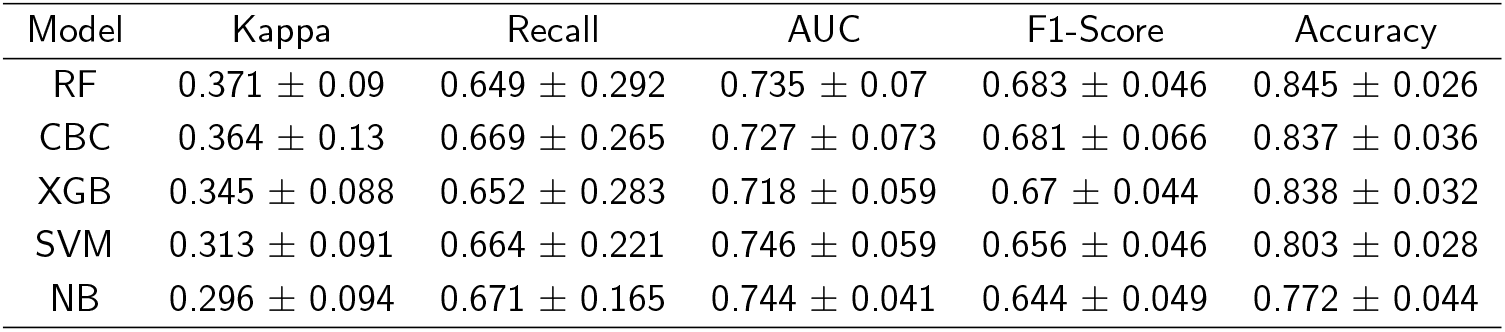
Top 5 models for the one-year death prediction outcome, obtained through cross-validation inside the primary 847 patients dataset. (Mean *±* SD)

| Model | Kappa | Recall | AUC | F1-Score | Accuracy |
| --- | --- | --- | --- | --- | --- |
| RF | 0.371 ± 0.09 | 0.649 ± 0.292 | 0.735 ± 0.07 | 0.683 ± 0.046 | 0.845 ± 0.026 |
| CBC | 0.364 ± 0.13 | 0.669 ± 0.265 | 0.727 ± 0.073 | 0.681 ± 0.066 | 0.837 ± 0.036 |
| XGB | 0.345 ± 0.088 | 0.652 ± 0.283 | 0.718 ± 0.059 | 0.67 ± 0.044 | 0.838 ± 0.032 |
| SVM | 0.313 ± 0.091 | 0.664 ± 0.221 | 0.746 ± 0.059 | 0.656 ± 0.046 | 0.803 ± 0.028 |
| NB | 0.296 ± 0.094 | 0.671 ± 0.165 | 0.744 ± 0.041 | 0.644 ± 0.049 | 0.772 ± 0.044 |

The data exploration process revealed the severe imbalance of 1:8, towards the negative result for 1 year death. However, this imbalance was not critical since there were still close to 100 patients representing the minority class and resampling techniques were viable in this binary classification setting. This outcome only makes use of 7 input variables (Table 5 - Appendix C), resulting from the feature selection process, and shows remarkably good results for such a small amount of input data.

The accuracy in the validation cohort was similar to that of the development cohort with an accuracy of 85% and an AUC of 0.74 (Appendix A).

**Table 5.**
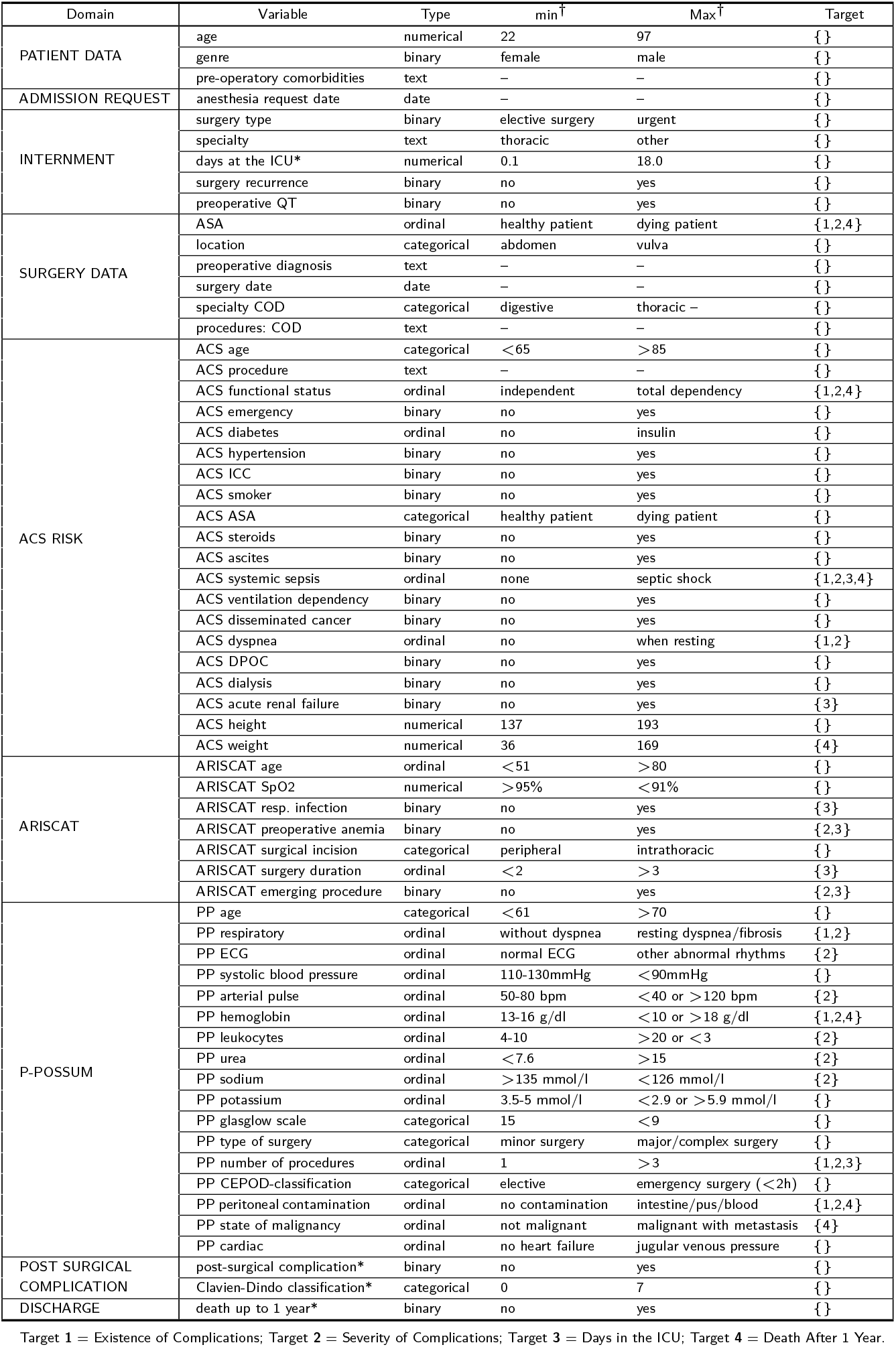
Summary of cohort data characteristics: input variables and output variables (star-marked). † Nominal values assumed to have an explicit ordering for value disclosure.

Previous studies have attempted to predict similar outcomes. Wang et al. [11] predicted 5-year mortality in a bladder cancer cohort of 117 patients with 0.8 accuracy, 0.86 sensitivity and 0.72 precision. Similarly, Corey et al. [12] included the prediction of 30-day mortality, with an AUC of 0.92, using information from 66,370 patients. Finally, Bihorac [25] predicted mortality for 1, 3, 6, 12 and 24 months after surgery with an AUC ranging from 0.83 for 1 month, to 0.77 for 24 months mortality. Furthermore, the 12 month outcome was predicted with an accuracy of 84%, a sensitivity of 0.38 and a specificity of 0.91. Although it is impossible to establish direct comparisons, due to distinct cohorts and study characteristics, our models offers potentially relevant results focusing the Portuguese population. Offering the possibility to predict 1-year death which might be a relevant information to complement medical prognosis on cancer postoperative survivability.

### Knowledge Extraction via Enhanced Associative Models

Given the competitive results of associative models, together with their unique knowledge extraction capabilities, further studies were conducted on these models. As an extension to the results obtained from this study, an improvement over traditional model representation is proposed.

The test set error is calculated for each node individually and displayed at leaf level. Additionally, leaf nodes are colored (according to the scheme at the top right corner of Fig. 3), traducing the error degree associated to the validation process.

**Figure 3.**
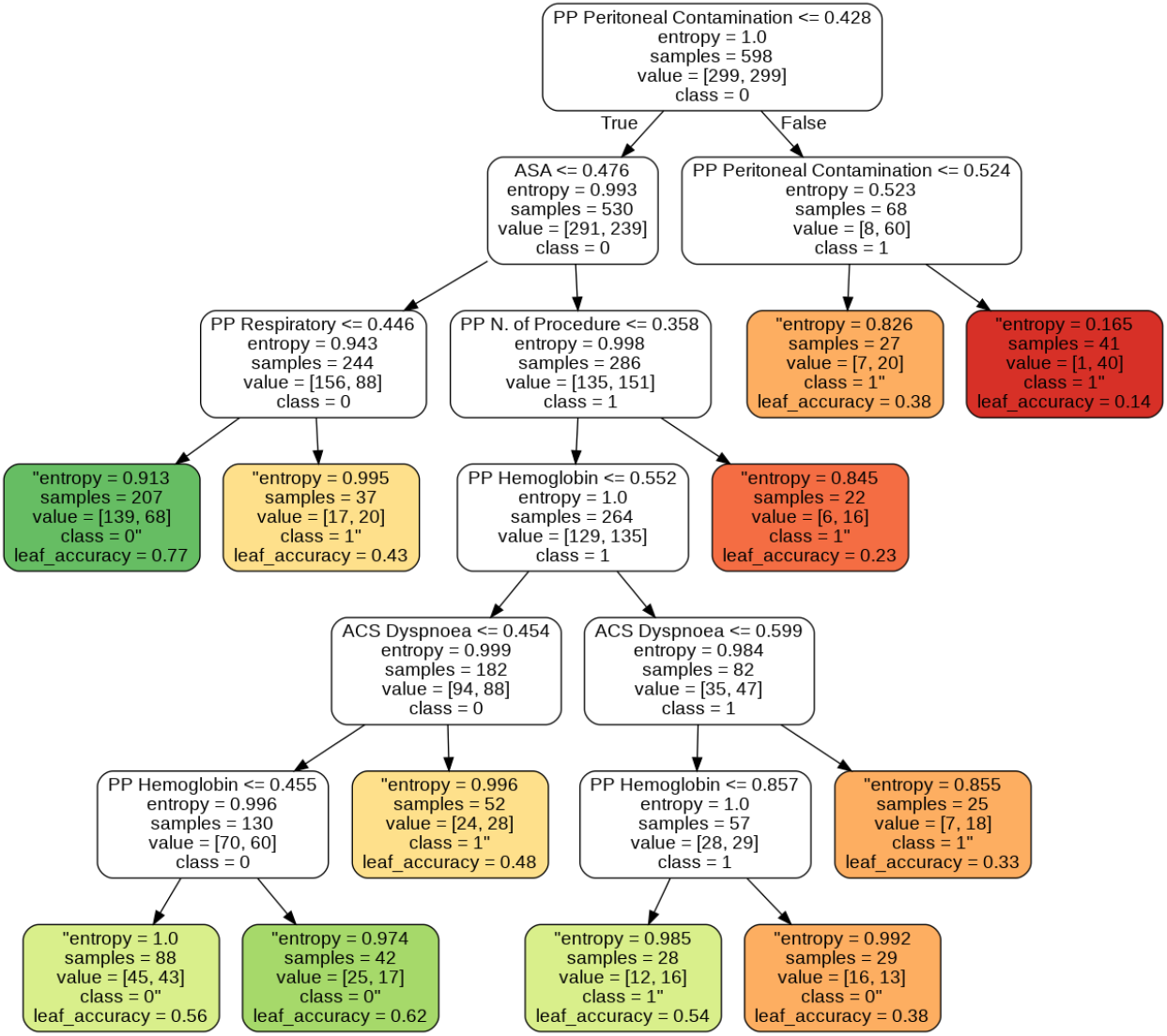
Example Decision Tree for the complication prediction outcome (green = better, red = worse).

This specific type of visualization is unmatched and can be further extended. Allowing for a quick assessment of the decision process, improving interpretability and confidence. This representation further helps doctors in the knowledge extraction process and in assessing the confidence level on the association rules captured by the models, and will eventually be implemented in tools used at the hospitals which contributed to the creation of this cohort study. An example graphical representation is presented in Fig.3, based on a Decision Tree used to predict the existence of complications. The full results for all outcomes are given as supplementary material available in an open repository (Appendix A).

### Variable Importance: Effort Allocation

Tree-based models not only stand out for their intuitive representation, but also for offering information about the importance of each feature in the prediction process. This information might be relevant for physicians in order to reduce the variable collection effort, that can reveal burdensome and too bureaucratic. Currently, IPO-Porto is collecting more than 60 pre-operative variables, but not all seem to be of paramount importance for the predictions. These models can indicate the relative feature importance for each input variable when making a prediction, contributing to their popularity in many areas, including clinical prognostication. A tool that is understandable and transparent contributes to an easier adoption and improved decision confidence. As an example, Fig. 4 gives the relative feature importance information relating to the prediction of the existence of complications, using DT, RF and XGBoost model. The full results for all outcomes are given as supplementary material in an open repository (Appendix A).

**Figure 4.**
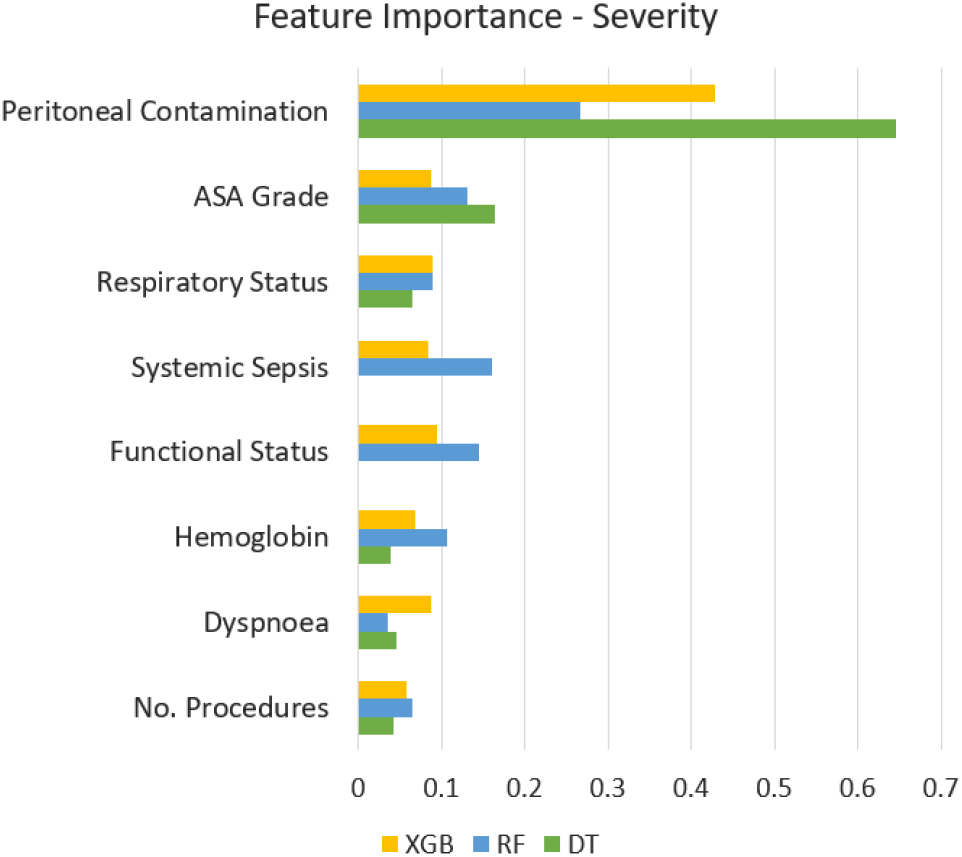
The 10 features with highest feature importance for the postoperative complication outcome in the XGB, RF and DT models.

## Clinical Decision Support System

Finally, we developed a web-based application to facilitate the usability of the selected models and to predict the four outcomes. The serialized predictive models can be used by clinicians in order to efficiently test cancer patients in preoperative time, after inserting the variables required for each outcome. The result provides the output of the models and the web-based score application is easy to use. Further explanations are delivered in the form of graphs. For the classification tasks, the predicted probabilities for the training set are plotted, as well as the probability for the current patient, to enable comparisons and further understand the confidence of the model. For the regression tasks, two graphs are plotted. One with the actual values versus the predicted values of the model, and a plot of the predictions’ residuals, both for the training data (see example in Figure 5). The web-based application and its code repository can be found in Appendix B.

**Figure 5.**
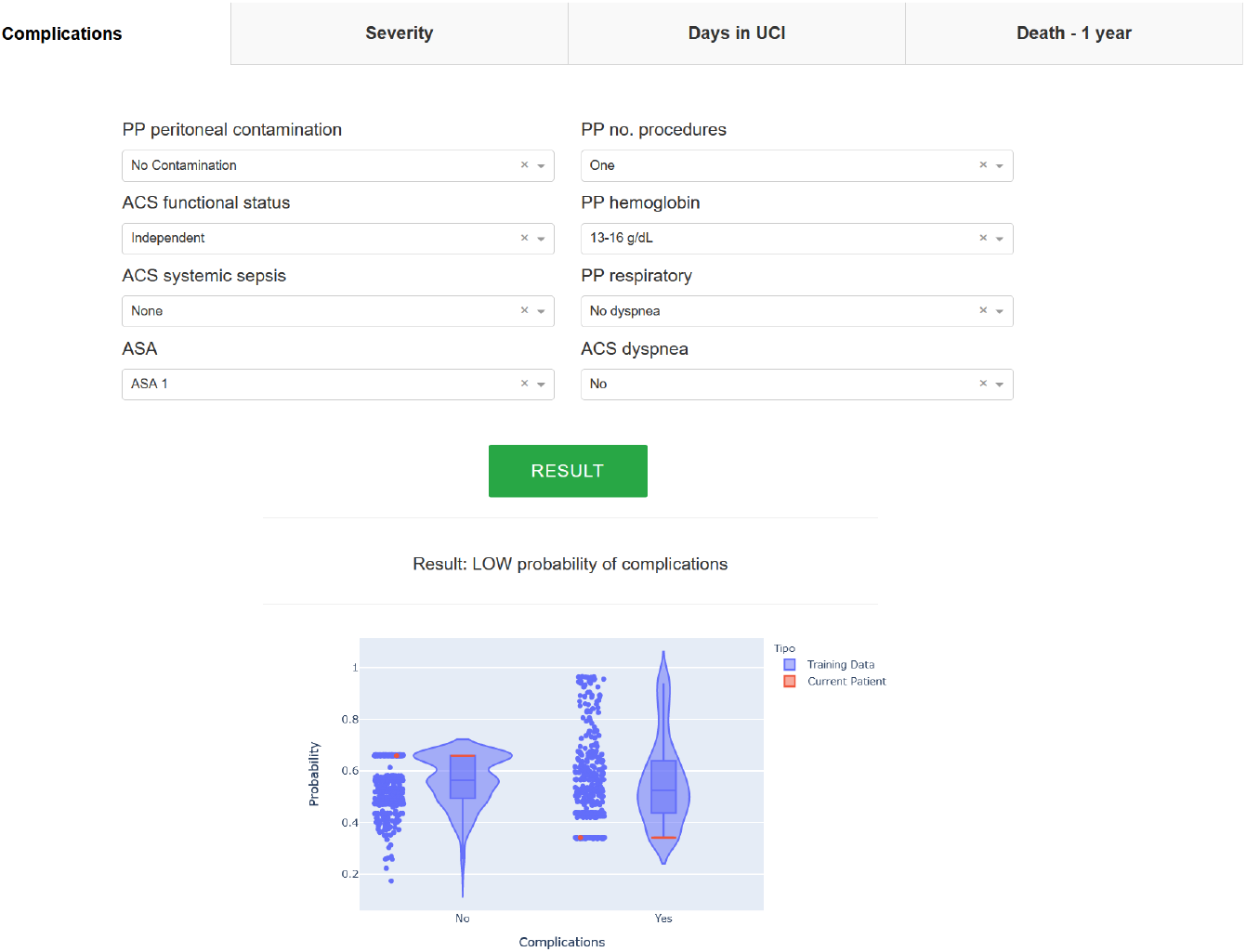
Screenshot example of the ‘Existence of Complications’ tab from the web application. The score includes 8 input fields and calculates a probability.

## Conclusions

In this work we have applied a machine learning approach for assessing the predictability of four major cancer surgical outcomes, with the goal of increasing the accuracy of previous traditional risk scores. We demonstrated that machine learning models derived from our local patient population were able to improve the accuracy of a previous traditional risk score. For these predictive models, we developed a web-based application based on variables measured at oncological hospitals, providing an improved approach for clinicians to estimate postoperative risk for cancer patients. Model interpretability is also enhanced, by offering new visualization options for tree-based models, in order to support medical decision processes. Additionally, information about relevant variables for the outcomes prediction is provided, contributing to more efficient data acquisition processes. Finally, the current implementation is being tested at IPO-Porto.

The main limitations of the present work are: i) missing values on the dataset, requiring imputation and possibly making the learning process more difficult, ii) difficulties on algorithms training due to the limited single-center cohort size.

In the future, IPO-Porto will also be releasing new datasets and extensions to already existing ones, which could impact the knowledge fed to the models improving them, especially in outcomes with severe imbalanced problems. External validation with data from other Portuguese cancer hospitals is also planned, which should further guarantee the generalization ability of these models.

## Data Availability

The data may be available upon a written request addressed to the authors.

https://github.com/danielmg97/cancer-prognostication-iposcore

## Competing interests

The authors declare that they have no competing interests.

## Acknowledgements

The authors thank IPO-Porto for providing the dataset. This work was supported by the FCT, through IDMEC, under LAETA project (UIDB/50022/2020), IPOscore project with reference DSAIPA/DS/0042/2018, and Data2Help (DSAIPA/DS/0044/2018). This work was further supported by the LAQV, financed by national funds from FCT/MCTES (UIDB/50006/2020) and the contract CEECIND/01399/2017.

## Appendix

## Appendix A Study Repository (Code and Results)

https://github.com/danielmg97/cancer-prognostication-iposcore

## Appendix B Beta Web App

Link: https://iposcore.herokuapp.com/

Repository: https://github.com/danielmg97/iposcore_webapp

## Appendix C Dataset Description

## Notes

### Competing Interest Statement

The authors have declared no competing interest.

### Author Declarations

The IPO-Porto Ethics Committee approved (CES IPO:91/019) the analysis of the anonymized data.

